# Feasibility and safety of renal denervation (RDN) in patients with end-stage renal disease (ESRD) and refractory hypertension (RHT)

**DOI:** 10.1101/2025.08.04.25332993

**Authors:** He Lingyun, Cai Han, Lai Yuxing, Zhou Wei, Su Jinzi, Fang Zhoufei

## Abstract

**Objective:** To evaluate the feasibility and safety of renal denervation (RDN) in patients with end-stage renal disease (ESRD) and refractory hypertension (RHT).

**Method:** A retrospective analysis was conducted on the baseline and follow-up data of ESRD patients who underwent RDN at the First Affiliated Hospital of Fujian Medical University between December 2017 and December 2024. Based on the etiology of ESRD, the patients were divided into three groups: diabetic nephropathy group (DN group), chronic glomerulonephritis group (CG group), and hypertensive nephrosclerosis group (HE group). RDN was performed using the NavStar pressure monitoring perfusion monopolar ablation catheter (Abbott, USA). A 3D model was constructed using the ENSITE system, and four-quadrant spiral point-by-point ablation was performed from distal to proximal on the main renal arteries and their branches bilaterally. Each ablation site was treated for 40 seconds at a temperature of 40°C. The ablation power was set at 8W for renal artery branches and 12W for the main branches.

**Result:** A total of 25 ESRD patients were enrolled, including 10 in the DN group, 8 in the CG group, and 7 in the HE group. All patients underwent regular hemodialysis with controlled dry weight. The number of ablation points was 15.04 ± 2.62 for the main branches and 4.68 ± 1.03 for the branch vessels. Fluoroscopy time was 23.64 ± 6.89 minutes, and contrast agent volume was 28.4 ± 8.5mL. No significant differences were observed among subgroups in terms of impedance, contrast agent volume, or fluoroscopy time before and after ablation. HE group had the least number of ablation points in both main and branch vessels. One case of vascular dissection requiring renal artery stent implantation occurred in the CG group, and one case of pseudoaneurysm occurred in the HE group. The median follow-up duration was 7.90 ± 1.55 months. During follow-up, one patient was lost to follow-up in each the DN and CG groups, and one patient in each the CG and HE groups died due to myocardial infarction. Major adverse cardiovascular events (MACE) occurred in one patient each in the CG and HE groups, both requiring hospitalization for heart failure. Postoperative follow-up was completed in 21 patients. Among them, 15 (71.43%) exhibited a positive response to RDN, including 8 (88.89%) in the DN group, 4 (66.67%) in the CG group, and 3 (50%) in the HE group. Compared with preoperative values, office systolic and diastolic blood pressure, as well as 24-hour ambulatory systolic and diastolic blood pressure, showed significant reductions after RDN.

**Conclusion:** Renal denervation (RDN) effectively reduced blood pressure in patients with end-stage renal disease (ESRD) and resistant hypertension (RHT). Among the etiological subgroups, patients with diabetic nephropathy may represent the most suitable candidates for RDN within the ESRD population.

## 1. BACKGROUND

End-stage renal disease (ESRD) represents the most severe stage of acute or chronic renal failure. In addition to disturbances in water, electrolyte, and acid-base balance, as well as dysregulation of renal endocrine functions, over 30% of ESRD patients develop refractory hypertension (RHT) [1], a prevalence significantly higher than that in the general hypertensive population. RHT is defined as the failure to achieve target blood pressure (below 140/90 mmHg) despite adherence to maximally tolerated doses of at least three antihypertensive agents, including a diuretic, for at least four weeks. Hypertension serves as both a cause and a critical driver in the onset and progression of chronic kidney disease (CKD). Notably, fewer than 20% of CKD patients attain adequate blood pressure control. RHT stands as a major risk factor for cardiovascular events and poor prognosis in CKD and ESRD patients [2, 3]. Consequently, effective blood pressure management remains an urgent clinical priority in CKD care.

ESRD and hypertension are interrelated, forming a vicious cycle. The pathogenesis of refractory hypertension (RHT) in ESRD is complex, primarily involving water-sodium retention, activation of the renin-angiotensin system (RAS), sympathetic overactivation, and dialysis-mediated clearance of antihypertensive drugs. Notably, RAS activation and sympathetic overexcitation play particularly significant roles [4,5]. During CKD progression, nephron loss and reduced renal blood flow directly and indirectly activate both the renin-angiotensin-aldosterone system (RAAS) and renal sympathetic nervous system. Active substances secreted by these systems, such as angiotensin and catecholamines, initially exert compensatory effects in early-stage CKD. However, excessive compensation exacerbates renal fibrosis, chronic inflammation, and cardiovascular damage, thereby accelerating CKD progression [5,6].Furthermore, sympathetic overactivation promotes hypertension through two mechanisms: 1) by increasing cardiac output and total peripheral resistance, and 2) by modulating central blood pressure regulation through its effects on vasomotor-controlling neurons in the brain [7]. As glomerular filtration rate declines, progressive enhancement of muscle sympathetic nerve activity can be observed in patients [8]. Particularly in hemodialysis-dependent ESRD patients, significant increases in nerve terminal density in renal arteries and perirenal tissues are evident, indicating heightened renal sympathetic activity compared to patients with CKD stages 1-4 [9].Animal studies of CKD have demonstrated that effective blockade of adrenergic receptors and suppression of sympathetic activity can inhibit the activation of pro-apoptotic, inflammatory and pro-fibrotic factors, thereby achieving both antihypertensive and renoprotective effects [10-12].

Renal denervation (RDN), an innovative interventional therapy in hypertension management, utilizes radiofrequency ablation to disrupt renal sympathetic nerve fibers. This procedure suppresses sympathetic overactivity and attenuates the blood pressure regulatory interaction between the kidneys and central nervous system, thereby achieving antihypertensive effects. Notably, current large-scale clinical trials on RDN uniformly exclude ESRD patients. However, several small-sample studies and case reports have demonstrated RDN’s efficacy in reducing blood pressure among ESRD patients [13-15]. The etiology of ESRD is complex, with the three primary causes being chronic glomerulonephritis, hypertensive nephrosclerosis, and diabetic nephropathy. The pathophysiological mechanisms underlying ESRD development vary across these etiologies, and the degree of sympathetic activation may likewise differ. Previous studies have not conducted subgroup analyses to determine which ESRD etiology responds most favorably to RDN.

## 2.1 METHODS

### 2.1. Study Design

This study enrolled 25 patients diagnosed with ESRD complicated by refractory hypertension (RHT) who underwent renal denervation (RDN) at the First Affiliated Hospital of Fujian Medical University and Shaowu Municiple Hospital of Fujian Province between December 2017 and December 2024. Baseline and follow-up evaluations included general demographic data, ambulatory blood pressure monitoring, antihypertensive medication regimens, hematological parameters, and echocardiographic indices. This is a registered clinical trial (Chinese Clinical Trial Registry identifier: ChiCTR-ONC-17012483 titled “Indication Screening and Technical Optimization of Renal Denervation for Refractory Hypertension.” The study protocol was approved by the Institutional Review Board of the First Affiliated Hospital of Fujian Medical University (Approval No.: 2017-001-02).

### 2.2. Inclusion and exclusion criteria

Inclusion and diagnostic criteria: (1) RHT diagnostic criteria: sufficient combination of three or more antihypertensive drugs for more than 4 weeks must include diuretics. The blood pressure in the third consultation room is ≥140/90mmHg, or the 24-hour ambulatory blood pressure is ≥ 135 mmHg; (2) Diagnostic criteria of 2)ESRD: The CKD definition and staging criteria revised by the Global Organization for Improving Prognosis of Kidney Disease in 2024 were adopted, that is, GFR < 15 ml/min; (3) The duration of dialysis in June ≤60 months, and the frequency of dialysis is 3 times a week; (4) The patient maintained dry weight within three months before RDN; (5) Renal artery images showed that the length of renal artery was ≥3cm and the diameter was ≥ 3 mm.

Exclusion criteria: (1) Unilateral renal and renal artery stenosis over 50%, renal tumor, and previous history of renal surgery or interventional surgery; (2) pregnant women or breastfeeding; (3) previously lost the ability of self-care due to severe atherosclerotic cardiovascular disease (ASCVD); (4) definite myocardial infarction, cerebrovascular accident, gastrointestinal bleeding and cirrhosis within 3 months; (5) Life expectancy is less than 1 year or diagnosed as malignant tumor; (6) Refuse to sign the informed consent.

### 2.3. Baseline Data

Clinical data from patients during baseline and follow-up periods were collected, primarily including: (1) General information: age, sex, height, dry weight, duration of hemodialysis, duration of hypertension, types of medications, blood pressure, and heart rate; (2) Creatinine, glomerular filtration rate, hemoglobin, albumin, uric acid, total cholesterol, triglycerides, low-density lipoprotein cholesterol, glycated hemoglobin, and urinary microalbumin;(4) Echocardiographic measurements: left atrial diameter, left ventricular end-diastolic diameter, interventricular septal thickness, and left ventricular ejection fraction; (5) 24-hour ambulatory blood pressure monitoring;(6) Major adverse cardiovascular events (MACE), including acute myocardial infarction, severe arrhythmia, heart failure, coronary heart disease-related death, stroke, and all-cause mortality;(7) Definition of the composite index of hypertension drug types and doses: the composite index was the weight of drug types × the total dose of drugs used. The weight of drug category was the type of drug used, 1 drug was defined as 1, 2 drugs were defined as 2, and so on. A standard dose of a drug was defined as 1, a half dose as 0.5, and a double dose as 2. The total dose of each drug was defined as the sum of the doses used.

### 2.4. Grouping method

According to the etiology of ESRD, they were divided into three groups, namely Diabetic Nephropathy group (DN group), chronic glomerulonephritis group (CG group) and hypertensive nephrosclerosis group (HE group). Some patients were divided into groups by renal biopsy in the early stage of the disease. If there was no renal biopsy, the patients would be grouped according to their medical history. DN group: (1) The course of diabetes was more than 10 years; (2) There was a test basis for moderate or large proteinuria in the early stage of the disease; (3) Blood pressure increased after renal function damage. CG group: (1) Proteinuria, hematuria, hypertension and edema were the basic clinical manifestations, and the course of disease was more than 10 years. HE group: (1) The course of hypertension was more than 10 years; (2) There was a test basis for microalbuminuria or oliguria in the early stage of the disease; (3) Renal dysfunction accompanied by hypertension.

### 2.5. implementation scheme of renal artery denervation

(1) After disinfection and towel spreading, 1% lidocaine was used for local anesthesia, and the patient’s right femoral artery was put into 8F vascular sheath, and 50IU/kg heparin was injected into the sheath; (2) Bilateral renal arteriography was performed through 6F JR3.5 contrast catheter; (3) Send the contact light-induced pressure unipolar ablation catheter of Abbott Company of America into bilateral renal arteries. Three-dimensional model was established by ENSITE system, and the catheter was ablated point by point from distal end to proximal end and from branch to main branch. Each site was ablated for 40s at 40°C. The ablation power of renal artery branch was 8w, and that of main branch was 12 W. (4) Re-examination of renal arteriography after ablation. (Figure 1)

**Figure 1.**
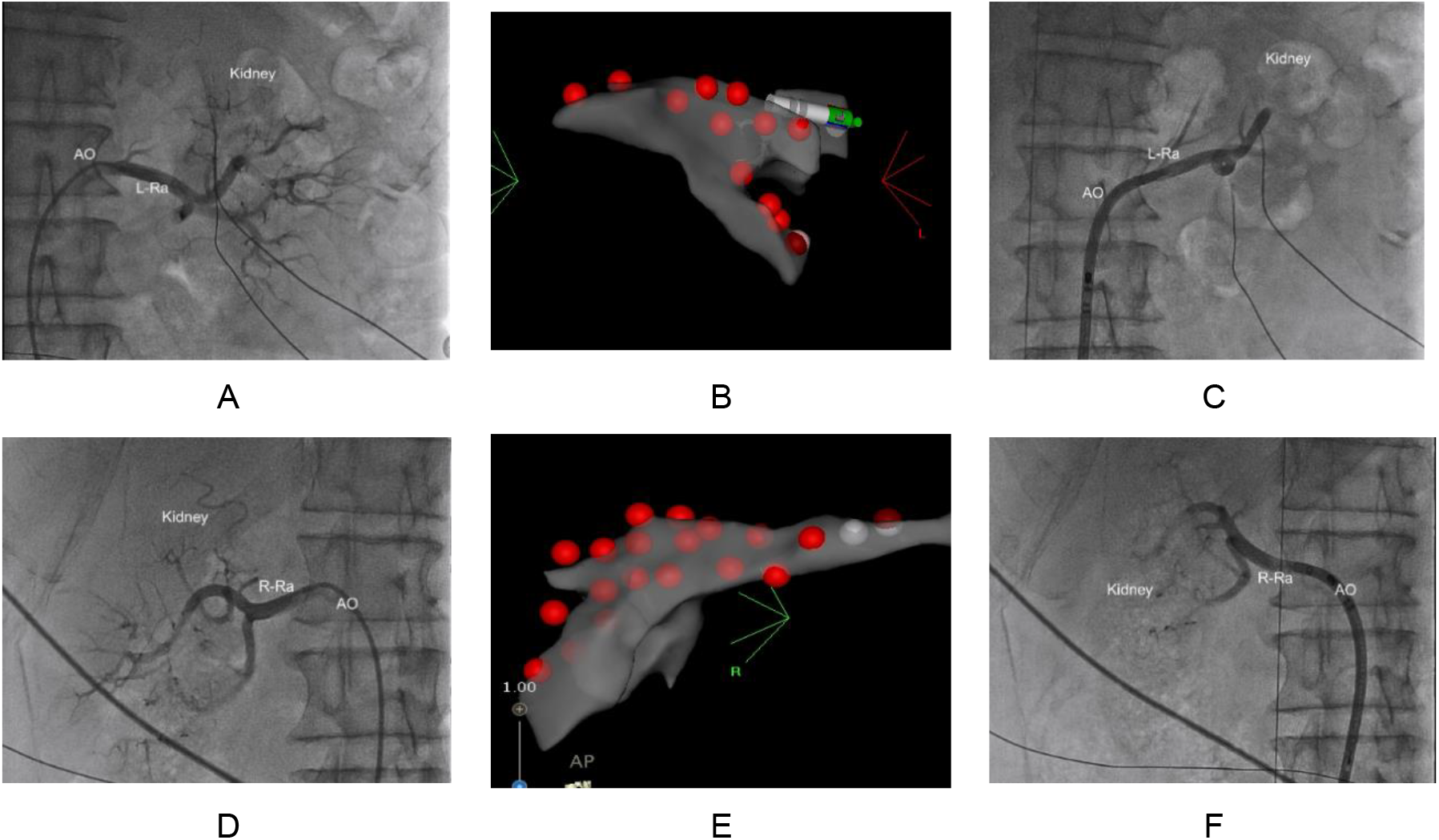
Procedure of renal artery denervation A: Left renal artery angiography before RDN; B: Three-dimensional modeling and point-by-point radiofrequency ablation of the left renal artery with a contact light-induced pressure unipolar ablation catheter; C: Postoperative left renal artery angiography; D: Preoperative right renal artery angiography; E: Three-dimensional modeling and point-by-point radiofrequency ablation of the right renal artery with a contact light-sensing pressure monopolar ablation catheter; F: Postoperative right renal artery angiography.

Important parameters such as ablation impedance, number of ablation points, contrast agent dosage and fluoroscopy time were recorded during operation. At the same time, the intraoperative safety parameters include: severe renal artery dissection affects blood flow and requires stent implantation, aortic dissection, femoral artery pseudoaneurysm or arteriovenous fistula. We reduce the blood pressure by more than 10mmHg or at least one drug type at the same blood pressure level, which was defined as a response to RDN.

### 2.6. Statistical analysis

Use SPSS 26.0 statistical software for statistical analysis. The measurement data of normal distribution was expressed by the mean standard deviation (SD), and the independent sample T test was used for comparison between two groups, and the ANOVA test was used for comparison between multiple groups. The measurement data of non-normal distribution was expressed by median (interquartile interval) [P50(P25, P75)], and nonparametric test was used for comparison between the two groups. Categorical data were expressed as number of cases (%), and intergroup comparisons were performed using the chi-square test. A two-tailed P-value < 0.05 was considered statistically significant.

## 3. RESULTS

### 3.1. Baseline data

A total of 25 patients were included in the study. There were 10 cases of ESRD due to diabetic nephropathy (7 males with an average age of 63.9 14.41 years), 8 cases of chronic glomerulonephritis (5 males with an average age of 53.00 9.58 years) and 7 cases of hypertensive nephropathy (5 males with an average age of 55.29 11.43 years). Glycosylated hemoglobin and fasting blood glucose in DN group were significantly higher than those in CG group and HE group. There was no significant difference in age, hemodialysis course, hypertension course, systolic and diastolic blood pressure in clinic, 24-hour ambulatory systolic and diastolic blood pressure, and compound indexes of antihypertensive drugs among the three groups. (Table 1)

**Table 1.** Baseline clinical data of the three groups.

|  | DN group (N=10) | CG group (N=8) | HE group (N=7) | P value |
| --- | --- | --- | --- | --- |
| Male (N/%) | 7 (70) | 6 (75) | 4 (57.14) | 0.749 |
| Age (yrs) | 63.9±14.41 | 53.00±9.58 | 55.29±11.43 | 0.159 |
| Dry weight (Kg) | 69±12.49 | 62.94±11.22 | 65.86±13.11 | 0.585 |
| Course of hypertension (yrs) | 10 (9.5,22.5) | 22.5 (12.5,30) | 20 (10,30) | 0.317 |
| The course of hemodialysis (yrs) | 3 (3,4) | 4 (3,4.75) | 3 (2,5) | 0.359 |
| ASCVD (N/%) | 7 (70) | 5 (62.5) | 5 (57.14) | 0.920 |
| Office systolic blood pressure (mmHg) | 163.50±27.83 | 163.75±10.28 | 150.29±9.20 | 0.321 |
| Office Diastolic blood pressure (mmHg) | 83.90±16.80 | 90.63±13.33 | 81.14±8.71 | 0.401 |
| Dynamic systolic blood pressure (mmHg) | 155.40±17.15 | 164.13±13.96 | 152.00±14.50 | 0.303 |
| Dynamic Diastolic blood pressure (mmHg) | 79.90±17.09 | 90.25±16.86 | 80.86±7.84 | 0.321 |
| Heart reat(bpm) | 70.60±10.98 | 72.13±6.42 | 69.86±13.95 | 0.915 |
| Hemoglobin (mmol/L) | 91.70±8.38 | 96.25±18.55 | 99.42±13.87 | 0.291 |
| Platelets (g/L) | 204.90±72.54 | 186.13±54.75 | 210.86±94.82 | 0.191 |
| Total bilirubin (μmol/L) | 6.28±2.12 | 6.43±4.20 | 5.94±3.07 | 0.557 |
| Albumin (g/L) | 34.97±6.16 | 36.7±4.58 | 36.77±6.24 | 0.651 |
| Cholesterol (mmol/L) | 3.63±0.93 | 3.69±0.69 | 3.47±0.68 | 0.323 |
| Triglyceride (mmol/L) | 1.34±0.63 | 1.62±0.37 | 1.04±0.60 | 0.050 |
| Low-density lipoprotein cholesterol (mmol/L) | 2.05±0.77 | 1.99±0.48 | 2.07±0.76 | 0.158 |
| High-density lipoprotein Cholesterol (mmol/L) | 0.99±0.49 | 1.01±0.45 | 1.02±0.47 | 0.937 |
| Glycated hemoglobin (%) | 8.66±2.69 | 5.34±0.53 | 5.03±0.34 | 0.009 |
| Fasting blood glucose (mmol/L) | 7.71±1.46 | 5.61±0.57 | 5.48±0.66 | 0.019 |
| Uric acid (mmol/L) | 466.87±126.32 | 393.10±102.75 | 455.58±103.31 | 0.946 |
| Creatinine (μmol/L) | 499.56±207.53 | 503.50±232.62 | 546.27±186.69 | 0.794 |
| Glomerular filtration rate (ml/min) | 11.34±5.08 | 12.28±6.08 | 11.29±4.66 | 0.379 |
| Serum potassium (mmol/L) | 4.48±0.73 | 3.91±0.63 | 4.52±1.19 | 0.730 |
| Serum sodium (mmol/L) | 141.14±3.87 | 142.19±4.63 | 142.06±5.97 | 0.206 |
| LAD(cm) | 4.39±0.45 | 4.65±0.52 | 4.73±0.42 | 0.439 |
| LVEDD(cm) | 5.15±0.56 | 5.64±0.46 | 5.55±0.29 | 0.173 |
| IVST(cm) | 1.37±0.25 | 1.51±0.39 | 1.55±0.42 | 0.697 |
| LVEF (%) | 60.93±5.24 | 59.93±7.67 | 63.48±6.79 | 0.466 |
| Drug composite index | 15.45±1.63 | 15.38±2.12 | 15.75±1.79 | 0.553 |

DN: Diabetic nephropathy; CG: Chronic glomerulonephritis; HE: Hypertensive nephrosclerosis; LAD: Left atrial diameter; LVEDD: Left ventricular end-diastolic diameter; IVST: Mitral valve thickness; LVEF: Left ventricular ejection fraction; ASCVD: Arteriosclerotic cardiovascular and cerebrovascular diseases.

### 3.2. Follow-up period data

The median follow-up time was 7.90±1.55 months. During the follow-up, one case in DN group and one case in CG group was lost. One case in CG group and one case in HE group died of myocardial infarction. There was one MACE event in CG group and one in HE group, both of whom were hospitalized due to heart failure. 21 patients were followed up after operation. Fifteen patients (71.43%) responded to RDN, including 8 patients (88.89%) in DN group, 4 patients (66.67%) in CG group and 3 patients (50%) in HE group (Figure 1). Compared with pre-operation, the systolic and diastolic blood pressure, 24-hour ambulatory systolic and diastolic blood pressure of 21 patients decreased significantly (Figure 2). Specific to each subgroup, compared with before operation, the systolic blood pressure, fasting blood glucose and IVST in DN group decreased significantly. The systolic blood pressure, diastolic blood pressure, 24-hour ambulatory systolic blood pressure and heart rate in CG group decreased significantly. Systolic blood pressure and diastolic blood pressure in HE group decreased significantly. (Table 2)

**Figure 1a.**
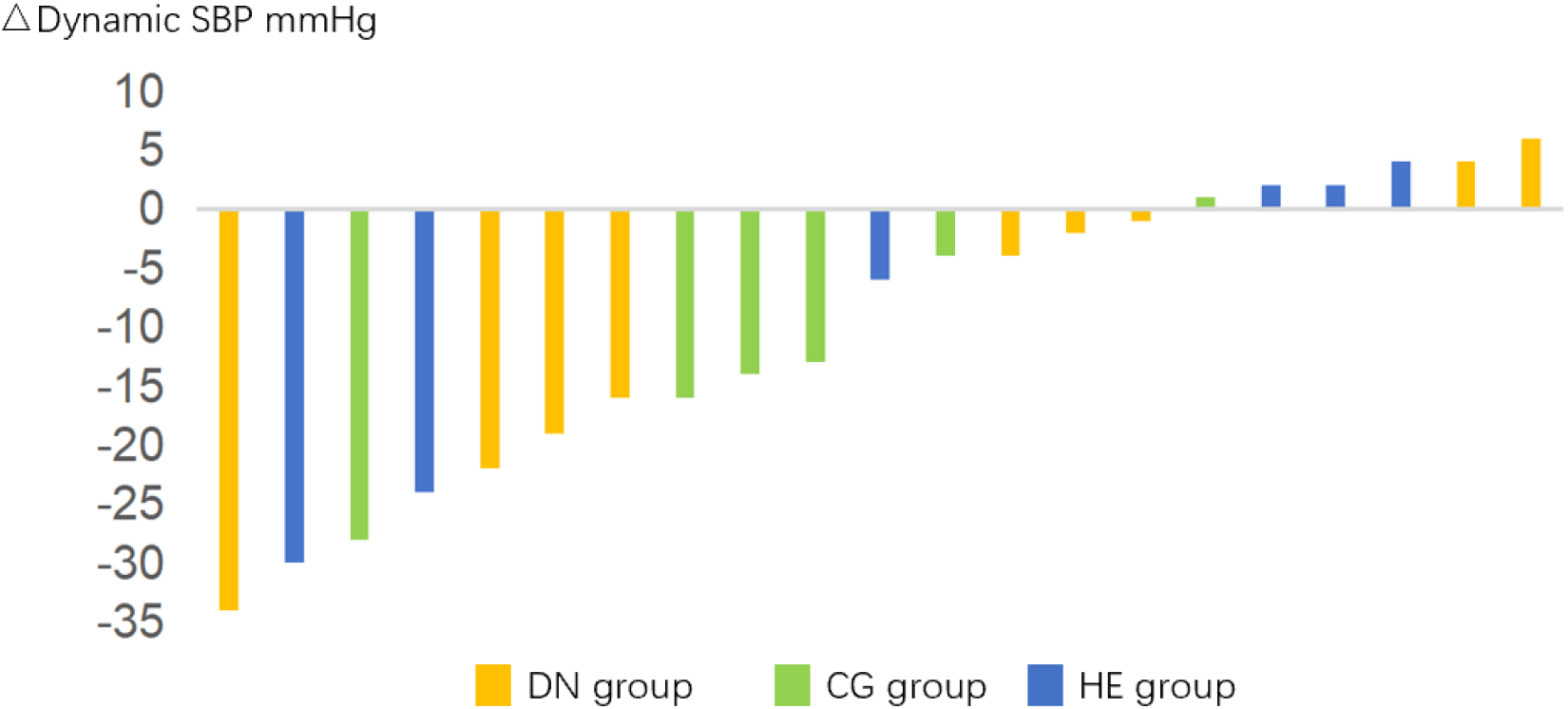
Response rate to RDN in the three groups

**Table 2.** Clinical data of patients during the follow-up period.

|  | DN group (N=9) | CG group (N=6) | HE group (N=6) | P value |
| --- | --- | --- | --- | --- |
| Average follow-up time (Months) | 7.78 ± 1.20 | 7.17 ± 1.33 | 8.83 ± 1.94 | 0.168 |
| Response rate of RDN | 8 (88.89%) | 4 (66.67%) | 3 (50%) | 0.457 |
| MACE | / | 1 (16.67) | 1 (16.67) | 0.437 |
| Office systolic blood pressure (mmHg) | 143.78±14.37* | 145.17±3.92* | 136.00±3.16* | 0.241 |
| Office Diastolic blood pressure (mmHg) | 80.56±8.88 | 79.83±12.29* | 76.67±3.67* | 0.705 |
| Dynamic systolic blood pressure (mmHg) | 144.78±9.97 | 146.50±2.67* | 141.17±6.74 | 0.480 |
| Dynamic Diastolic blood pressure (mmHg) | 76.78±11.05 | 80.83±11.48 | 78.00±5.80 | 0.745 |
| Heart reat(bpm) | 70.11±3.89 | 68.67±5.09* | 66.83±8.52 | 0.576 |
| Hemoglobin (mmol/L) | 94.33±7.35 | 96.00±14.09 | 103.33±12.37 | 0.465 |
| Platelets (g/L) | 192.44±24.79 | 195.33±39.25 | 216.67±66.10 | 0.037 |
| Total bilirubin (μmol/L) | 6.83±2.02 | 6.65±2.56 | 5.87±1.62 | 0.350 |
| Albumin (g/L) | 33.46±3.53 | 35.35±2.98 | 36.18±2.64 | 0.824 |
| Cholesterol (mmol/L) | 3.97±0.65 | 3.60±0.76 | 3.52±0.46 | 0.610 |
| Triglyceride (mmol/L) | 1.34±0.63 | 1.62±0.37 | 1.04±0.59 | 0.015 |
| Low-density lipoprotein cholesterol (mmol/L) | 2.26±0.55 | 1.93±0.59 | 2.22±0.32 | 0.344 |
| High-density lipoprotein Cholesterol (mmol/L) | 0.90±0.29 | 0.98±0.34 | 0.99±0.31 | 0.836 |
| Glycated hemoglobin (%) | 7.51±0.86 | 5.53±0.34 | 5.15±0.34 | 0.003 |
| Fasting blood glucose (mmol/L) | 6.92±0.69* | 5.65±0.39 | 5.18±0.34 | 0.061 |
| Uric acid (mmol/L) | 436.78±70.11 | 422.50±44.01 | 438.05±99.24 | 0.167 |
| Creatinine (μmol/L) | 517.33±225.42 | 414.44±160.40 | 560.00±187.94 | 0.362 |
| Glomerular filtration rate (ml/min) | 11.43±4.40 | 14.18±5.34 | 11.13±4.83 | 0.824 |
| Serum potassium<br>(mmol/L) | 4.41±0.37 | 4.33±0.68 | 4.42±0.54 | 0.277 |
| Serum sodium<br>(mmol/L) | 140.67±2.23 | 142.00±3.22 | 138.33±2.58 | 0.970 |
| LAD (cm) | 4.40±0.40 | 4.44±0.44 | 4.87±0.34 | 0.996 |
| LVEDD (cm) | 5.09±0.36 | 5.53±0.43 | 5.40±0.19 | 0.152 |
| IVST (cm) | 1.29±0.22* | 1.51±0.36 | 1.47±0.30 | 0.524 |
| LVEF (%) | 59.78±3.42 | 58.83±6.74 | 62.17±6.01 | 0.200 |
| Drug composite<br>index | 14.33±1.41 | 14.00±0.89 | 14.67±0.52 | 0.135 |

\* significant changes compared to before RDN, $P < 0.05$ ; DN: Diabetic nephropathy; CG: Chronic glomerulonephritis; HE: Hypertensive nephrosclerosis; LAD: Left atrial diameter; LVEDD: Left ventricular end-diastolic diameter; IVST: Mitral valve thickness; LVEF: Left ventricular ejection fraction; MACE: Major adverse cardiovascular events.

**Figure 2.**
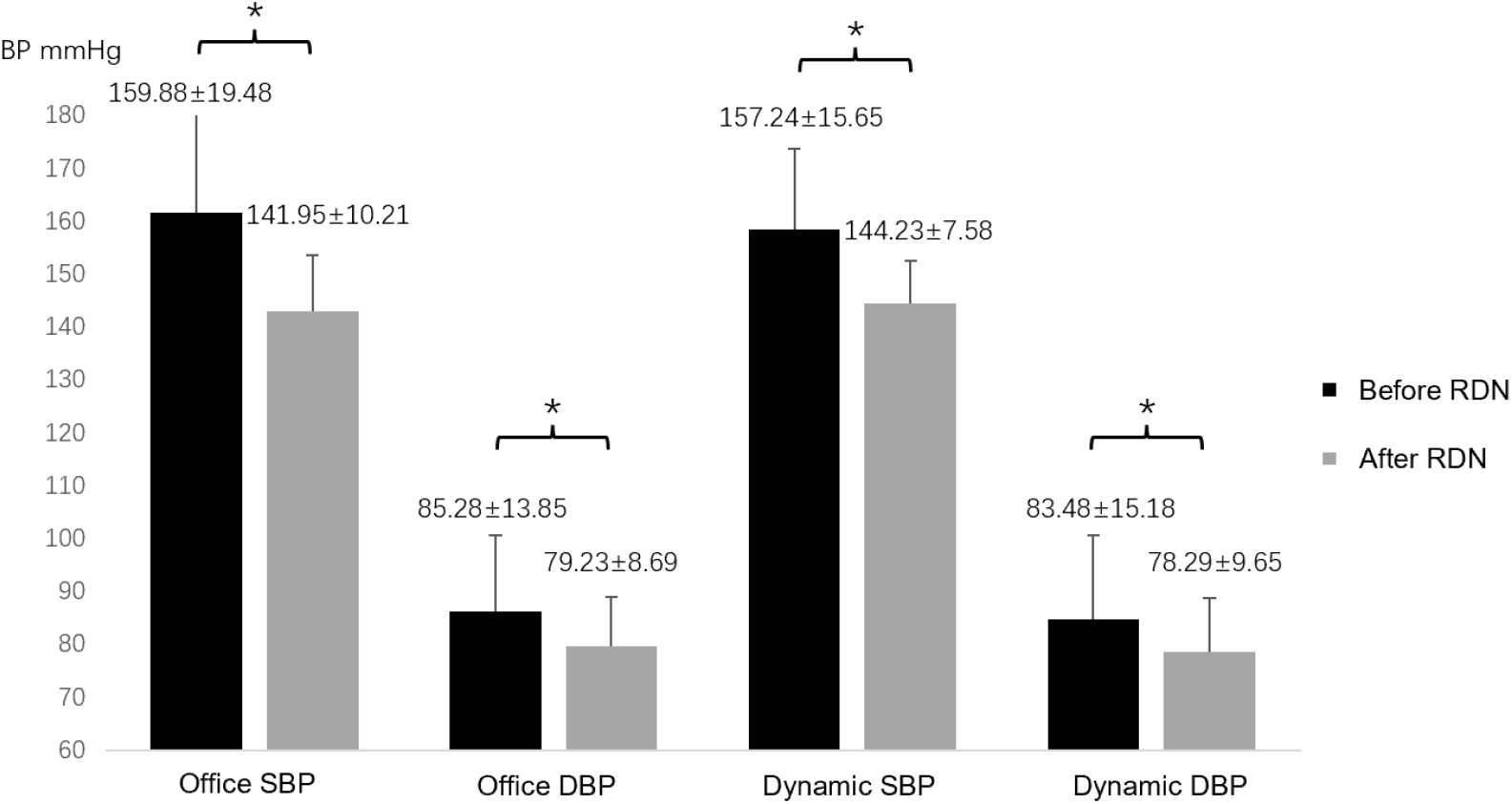
Changes in office and dynamic blood pressure before and after the RDN procedure

### 3.3. RDN parameters and complications

In 25 patients, the main branch ablation points were 15.04±2.62, the branch ablation points were 4.68±1.03, the fluoroscopy time was 23.64±6.89min, and the contrast agent dosage was 28.4±8.5ml. In each subgroup, the long diameter and short diameter of kidney in HE group were the shortest, and the ablation points of main branch and branch were the least. There was no significant difference in impedance, contrast agent dosage and fluoroscopy time before and after ablation in each subgroup. In CG group, 1 case of vascular dissection was implanted with renal artery stent, and in HE group, 1 case of pseudoaneurysm occurred. (Table 3)

**Table 3.**
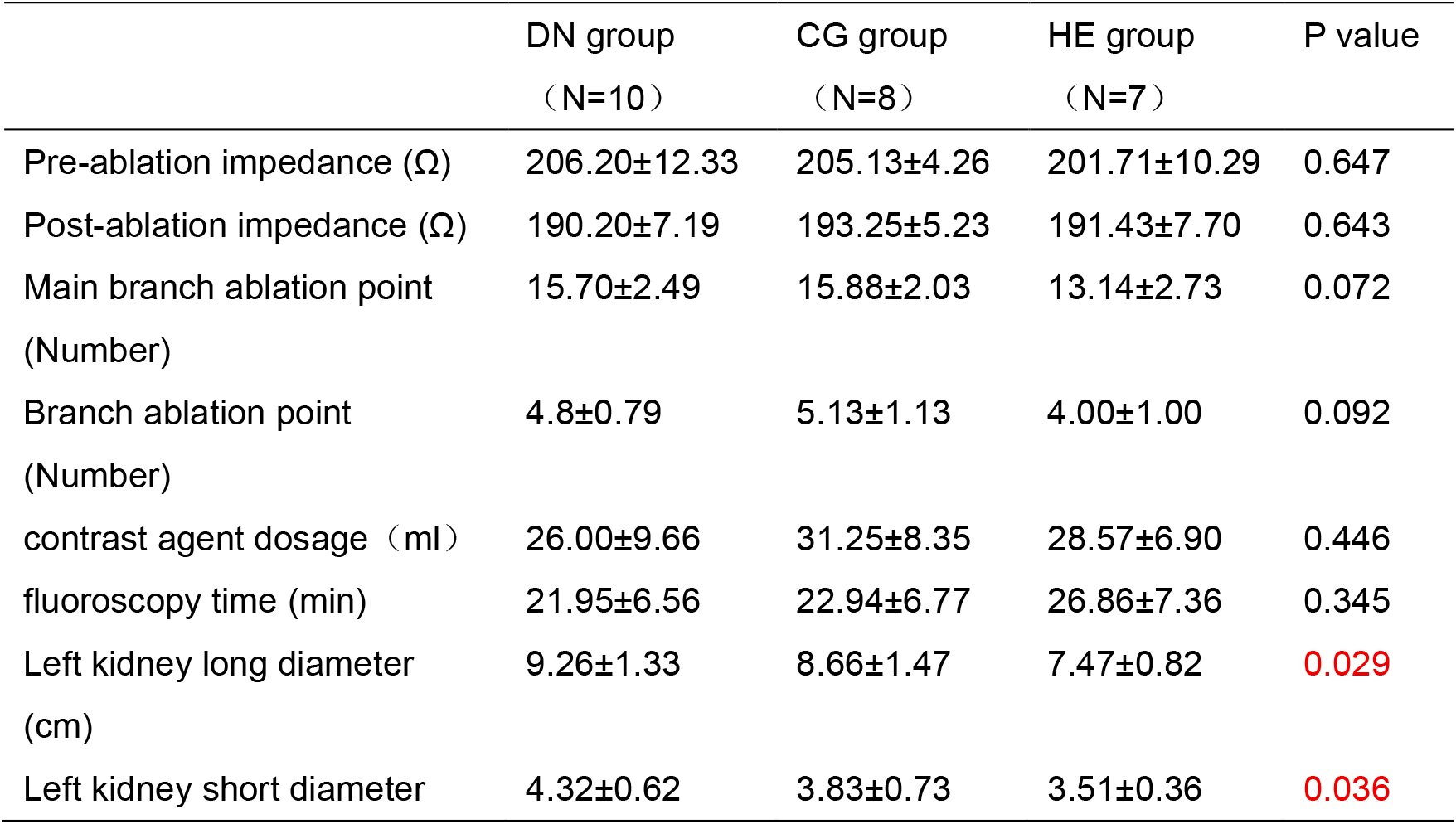
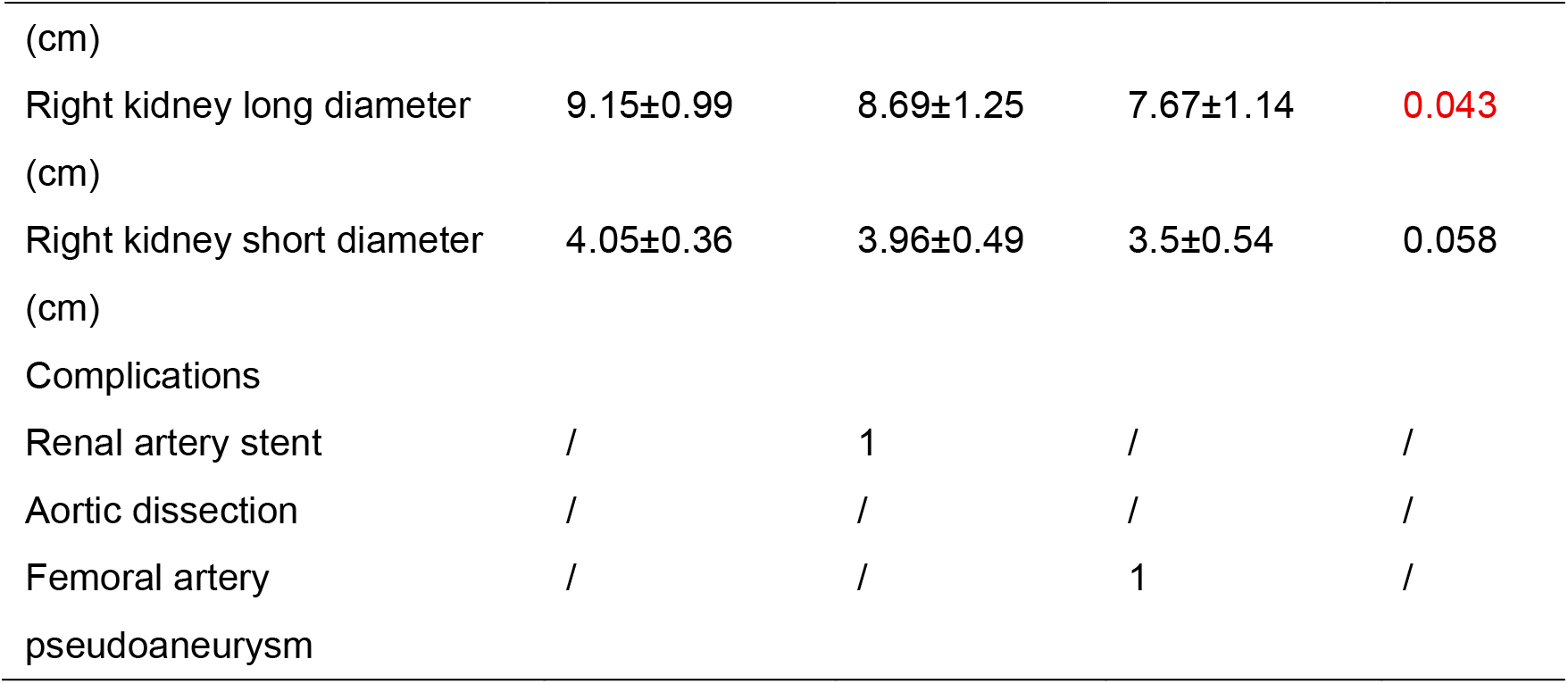
RDN parameters and complications.

## 4. DISCUSSION

According to the 2021 KDIGO guidelines [16], the target systolic blood pressure (SBP) for CKD patients was set below 120 mmHg. A meta-analysis of data from 1 million adult hypertensive individuals demonstrated that lowering the SBP target to below 120 mmHg in CKD patients with hypertension significantly reduces the incidence of cardiovascular events and all-cause mortality [17]. Therefore, it is essential to recognize the necessity of intensive blood pressure management to further lower blood pressure targets in CKD patients. The benefits of blood pressure reduction were evident, with every 10 mmHg decrease in SBP proven to reduce the incidence of coronary heart disease, stroke, and heart failure [18]. Particularly for CKD patients, especially those with ESRD, lowering blood pressure can reduce ASCVD risk. Maintaining dry weight through regular hemodialysis was a prerequisite for blood pressure control in these patients. Current common treatments for ESRD with resistant hypertension (RHT) include increasing peritoneal dialysis, high-flux dialysis, low-sodium dialysis, or combining aldosterone receptor antagonists with angiotensin-converting enzyme inhibitors on top of existing medications. These methods have been shown to lower blood pressure in some ESRD patients [19-20]. In terms of interventional therapies, renal artery embolization, nephrectomy, and renal artery stenting are suitable for specific types of ESRD patients, such as those with massive proteinuria, renal tumors, or severe renal artery stenosis [21-23].This study confirmed that renal denervation (RDN) can be used to treat ESRD patients with RHT. Despite disuse atrophy of renal arteries and kidneys in many patients, which reduced ablation sites, the response rate to RDN still exceeds 70%. Therefore, RDN can be considered a new therapeutic option for blood pressure management in this patient population.

Not all patients with resistant hypertension (RHT) were suitable candidates for renal denervation (RDN). Evidence suggested that younger patients, those with tachycardia, elevated plasma renin levels, increased blood pressure variability, obesity, those who had undergone accessory renal artery treatment, and those with less aortic calcification tended to respond better to RDN in terms of blood pressure reduction [25-27]. This indicated that patients with heightened sympathetic nervous system activity might have had a higher response rate to RDN. However, there was currently a lack of widely available, simple, and routine tests to assess sympathetic excitability. Therefore, we could only explore predictive factors for a favorable RDN response based on etiology and disease characteristics. Prospective clinical studies also demonstrated that RDN significantly reduced local renal sympathetic activity by 47% [28]. The results of this study confirmed that RDN in ESRD patients with RHT led to a significant postoperative reduction in blood pressure. Among ESRD patients with different underlying causes, those with diabetic nephropathy exhibited the greatest blood pressure reduction.

The renal sympathetic nervous system affects blood pressure through two pathways. It supplies the kidneys through output fibers located in the adventitia of the renal artery and the sympathetic nerve network. It also transmits signals back to the central nervous system through incoming sympathetic nerve fibers in the adventitia. Stimulation of the renal efferent sympathetic nerve increases the reabsorption of water and sodium ions by the renal tubules and the constriction of renal vessels. Decreased renal blood flow and glomerular filtration lead to an increase in renin and norepinephrine secretion, causing an increase in blood pressure. In this study, the HE group showed the smallest renal length and width, suggesting that patients with hypertensive nephrosclerosis developed more severe renal atrophy compared to the other two groups. Renal artery angiography demonstrated particularly sparse primary and secondary branches of the renal artery, indicating an ischemic state of the renal microcirculation.This microcirculatory ischemia could not be alleviated by RDN. The persistent renal ischemia directly stimulated renal afferent sympathetic nerves, which subsequently affected hypothalamic activity and led to increased secretion of norepinephrine and angiotensin I. Previous studies had established that afferent nerves played a critical role in mediating the blood pressure-lowering effects of renal denervation [29].Furthermore, the sparse branching pattern of the renal arteries directly limited the number of available ablation sites in the branches. Published research had confirmed that a combined ablation approach targeting both the main trunk and branches yielded better outcomes than ablation of the main trunk alone [30]. These pathophysiological and anatomical factors likely contributed to the lower response rate to RDN observed in patients with hypertensive nephrosclerosis compared to the other two groups.

Muscle sympathetic nerve activity (MSNA) was demonstrated to induce hypertension and was associated with impaired insulin sensitivity. Hyperinsulinemia, known as an etiological factor of type 2 diabetes mellitus (T2DM), was shown to promote sympathetic activation [31-32]. Previous studies indicated that even after excluding confounding factors including obesity, hypertension and metabolic syndrome, sympathetic activation persisted in T2DM patients [33-35]. Moreover, compared with patients having impaired glucose tolerance, T2DM patients exhibited increased sympathetic excitability and elevated norepinephrine spillover, suggesting progressive enhancement of sympathetic activity with disease advancement [36]. Based on these theoretical foundations, current research has been conducted to evaluate the therapeutic efficacy of RDN in metabolic cardiovascular diseases. Diabetic nephropathy, being a common complication of diabetes and a leading cause of mortality among diabetic patients, was specifically investigated in this study. Our results confirmed an excellent response rate (88.89%) of RDN in end-stage diabetic nephropathy patients. This remarkable therapeutic effect would undoubtedly contribute to reducing ASCVD events in this patient population.

Chronic glomerulonephritis (CG) was found to induce hypertension through multiple mechanisms, including activation of the RAAS system, uremic toxin accumulation, impaired baroreflex function, endothelial dysfunction, and excessive sympathetic activation. Among these, RAAS activation and excessive sympathetic activation were identified as the predominant mechanisms underlying hypertension development in CG patients. Animal studies demonstrated that CG mouse models exhibited a vicious “renal-brain neural” circuit: renal afferent nerves → subfornical organ → paraventricular nucleus → rostral ventrolateral medulla → spinal cord (T9-T11) → renal efferent nerves. When renal afferent nerves became excessively activated, this renal-brain circuit showed hyperactivity, leading to overexcitation of renal sympathetic nerves. The excessive activation of renal sympathetic nerves was shown to increase RAAS activity, elevate blood pressure in CG mice, and exacerbate cardiac and renal structural abnormalities along with dysfunction [37]. In CG models, various pro-inflammatory factors including IL-1β, IL-2, IL-6, and TNF-α were observed to participate in immune response regulation, causing vascular injury and endothelial dysfunction that ultimately resulted in elevated blood pressure. Through inflammatory processes, CG was found to induce excessive activation of the sympathetic nervous system while impairing vascular endothelial function, leading to progressively worsening hypertension, deteriorating renal function and structure, and creating treatment-resistant hypertension [38].Research indicated that renal denervation (RDN) could reduce angiotensin II-NADPH oxidase-mediated oxidative stress by blocking procalcitonin receptors or α2-adrenergic receptors. This intervention was shown to decrease macrophage infiltration in CG models and inhibit the activation of inflammatory molecules and pro-inflammatory factors. Therefore, for CG patients, RDN appeared to interrupt sympathetic overactivity caused by inflammation, thereby achieving blood pressure control [39].This study confirmed that RDN had a clear antihypertensive effect in CG patients. Furthermore, the intervention showed potential to somewhat alleviate renal function deterioration in these patients, reduce cardiovascular events within this population, and delay progression to end-stage renal disease.

The present study had several limitations, including its single-center design, small sample size, and lack of a sham procedure control group. As a pilot investigation, this study laid the groundwork for future multicenter, large-scale clinical trials with expanded patient enrollment. In subsequent research phases, we plan to both increase the overall sample size and incorporate a more diverse population of ESRD patients with varying etiologies. This expanded approach will enable more comprehensive evaluation of the specific patient characteristics that predict optimal responses to RDN therapy in the ESRD population.

## 5. CONCLUSION

Our study demonstrated the progress toward precision medicine in RDN. The results confirmed that RDN exerted a clear antihypertensive effect in patients with ESRD and RHT, with both feasibility and safety being well validated. Furthermore, it was notably observed that ESRD patients with diabetic nephropathy as the underlying etiology exhibited the highest response rate to RDN. This research expanded the potential applications of RDN and provided evidence to support its use in treating CKD and ESRD patients.

## AUTHOR DECLARATIONS

The authors have no conflicts to disclose.

## ETHICS APPROVAL DECLARATIONS

This is a registered clinical trial (Chinese Clinical Trial Registry identifier: ChiCTR-ONC-17012483 titled “Indication Screening and Technical Optimization of Renal Denervation for Refractory Hypertension.” The study protocol was approved by the Institutional Review Board of the First Affiliated Hospital of Fujian Medical University (Approval No.: 2017-001-02).

## INFORMED CONSENT FOR INVASIVE OPERATIONS

The patient has signed an informed consent prior to RDN and echocardiography, and is fully aware of the risks and benefits of the examination, and is performed by a qualified physician.

## PRIVACY PROTECTION CONFIRMATION

The patient’s personal information has been anonymized and there is no identifiable information in the article.

## AUTHOR CONTRIBUTIONS

Zhoufei Fang contributed to conception and design of the study. Han Cai, Luxing Lai and Lingyun He performed the RDN and data collection. Wei Zhou and Zhoufei Fang contributed to interpretation and manuscript writing and performed the data analysis. Also, she is responsible for English writing and expression. All authors approved the final manuscript.

**Lingyun He**: Data curation (equal); Formal analysis (equal); Investigation (equal); Methodology (equal); Writing – original draft (equal); Writing – review & editing (equal).

**Zhoufei Fang:** Data curation (equal); Formal analysis (equal); Investigation (equal); Methodology (equal); Writing – original draft (equal); Writing – review & editing (equal).

**Han Cai:** Conceptualization (equal); Funding acquisition (equal); Supervision (equal); Writing – original draft (equal); Writing – review & editing (equal).

## DATA AVAILABILITY

The data that support the findings of this study are available from the corresponding author upon reasonable request.

## SIGNIFICANCE STATEMENT

This paper focuses on the feasibility and safety of renal denervation (RDN) in patients with end-stage renal disease (ESRD) and refractory hypertension (RHT). The main purpose of this discussion is to explore which type of ESRD patients would benefit more from RDN and to analyze the possible reasons for this. By analyzing the blood pressure of ESRD patients and their response rate to RDN, it is proved that RDN effectively reduced blood pressure in patients with ESRD and RHT. Among the etiological subgroups, patients with diabetic nephropathy may represent the most suitable candidates for RDN within the ESRD population.

